# Shared Genetic Architecture and Neurobiological Pathways of Problematic Alcohol Use and Anxiety Disorders

**DOI:** 10.1101/2025.11.24.25339061

**Authors:** Mingjian Shi, Tommy Gunawan, Samantha G. Malone, Michael Setzer, Zachary Piserchia, Christal N. Davis, Christopher T. Rentsch, Eleni Friligkou, Hang Zhou, Renato Polimanti, Lu Wang, Nataraja Sarma Vaitinadin, Henry R. Kranzler, Joshua C. Gray

## Abstract

**Background:** Problematic alcohol use (PAU) and anxiety disorders (ANX) frequently co-occur, implying shared genetic and neurobiological foundations. However, the directionality of potential causal relationships and the specific mechanisms underlying the overlap remain unclear. Thus, we investigated the shared genetic architecture and neurobiological pathways between PAU and ANX using a multimethod genomic approach.

**Methods:** We analyzed summary statistics from genome-wide association studies (GWAS) of PAU and ANX using Mendelian randomization to assess causal associations between ANX and PAU. We used MiXeR to assess the overall shared genomic architecture, Local Analysis of (co)Variant Association to estimate regional genetic correlations, and conjunctional false discovery rate (conjFDR) to identify individual overlapping loci. We used FUMA to map single-nucleotide polymorphisms (SNPs) to independent loci, conduct differential gene expression analyses across 30 general and 54 specific tissue types, and perform cell-type specificity analyses using a human brain cell atlas. Druggability of identified targets was also evaluated.

**Results:** Mendelian randomization analyses indicated bidirectional causal associations between ANX and PAU. MiXeR identified moderate polygenic overlap (52.5%) and genetic correlation (*r_g_* = 0.44) between the traits, with high effect direction concordance among shared variants (86.4%). ConjFDR identified 97 shared lead SNPs, of which 89 had concordant and 8 discordant effects on PAU and ANX. These loci mapped to 97 genes, including *DRD2* and *PDE4B*, genes linked to dopaminergic and cAMP signaling pathways, respectively. Concordant gene expression was enriched in brain, nerve, adrenal gland, esophagus, stomach and colon, with enriched expression specifically in the prefrontal cortex, anterior cingulate cortex, hippocampus, hypothalamus, substantia nigra and amygdala. FUMA cell-type enrichment analysis identified associations predominantly in neurons from the cerebral cortex, hippocampus, and thalamus.

**Conclusions:** We found substantial genetic and neurobiological overlap of PAU and ANX, highlighting reciprocal, causal relationships between the traits, with differentially expressed genes enriched in addiction- and anxiety-relevant brain regions. These findings support shared underlying causal mechanisms linking the two conditions.

## Introduction

Twenty to 40% of individuals with anxiety disorders (ANX) are also diagnosed with alcohol use disorder (AUD) (Castillo-Carniglia et al., 2019; Lai et al., 2015). Moreover, individuals with ANX and depression experience more AUD symptoms than controls who drink at the same level (Anker et al., 2023). The frequent co-occurrence of AUD and ANX is thought to stem from shared etiology (i.e., genetics and pathophysiology) and reciprocal physiological effects. For example, drinking alcohol is acutely anxiolytic and frequently serves as a coping or self-medicating mechanism in ANX, increasing risk for AUD and problematic drinking behaviors among individuals with ANX (Anker & Kushner, 2019; Goodman et al., 2022). Furthermore, individuals with ANX can transition rapidly from binge drinking and intoxication to dependence, where intensified negative affect and withdrawal occur during abstinence (Anker & Kushner, 2019). Individuals with ANX also experience more negative consequences when drinking (Napper et al., 2015) and may experience more hangover anxiety after drinking (Kim et al., 2023).

Recent advances in psychiatric genetics have begun to clarify the biological relationship between ANX and problematic alcohol use (PAU; i.e., alcohol use disorder, alcohol dependence, and alcohol-related problems measured by the Alcohol Use Disorders Identification Test—Problems score [AUDIT–P]). For example, a meta-analysis of generalized anxiety disorder, panic disorder, social phobia, agoraphobia, and specific phobias across three GWAS showed a moderate positive genetic correlation between ANX and PAU (Colbert et al., 2021). The recent release of a large, multi-ancestry GWAS of ANX (Friligkou et al., 2024) provides unprecedented statistical power to interrogate these links. Coupled with analytic approaches such as Mendelian randomization (MR) to test causal directions, MiXeR to quantify shared polygenicity, and conjFDR and FUMA to pinpoint overlapping loci and their functional consequences, it is now possible to move beyond correlation to obtain a more detailed understanding of the shared genetic architecture and neurobiological pathways connecting ANX and PAU. Clarifying the biological systems that drive their overlap could make it possible to identify novel therapeutic targets for comorbid ANX and PAU.

Prior work using summary statistics from large GWAS has elucidated the overlapping genetic and neurobiological foundations of psychiatric disorders and related traits (e.g., Hindley et al., 2022; Hope et al., 2023; Tesfaye et al., 2023). For instance, we recently identified overlapping genetic variants associated with body mass index (BMI) and AUD (Malone et al., 2025), which showed that shared loci mapped to key brain regions implicated in both eating behavior and addiction. Here, we extend this approach to systematically investigate the causal relations and shared genetic architecture of PAU and ANX, integrating methods to estimate directionality, quantify polygenic overlap, localize shared loci, and functionally annotate their downstream expression patterns.

## Methods

### Genome-wide association studies

We utilized summary statistics based on the GWASs for ANX (Friligkou et al., 2024) and PAU (Zhou et al., 2023) with modified cohorts to minimize sample overlap. The European ancestry (EUR) subset for ANX included a meta-analysis of 107,415 from the All of Us Research Program (AoU) and 418,856 from FinnGen biobank Release 12 (Figure S1). For ANX multi-ancestry (MA) cohorts, we performed a meta-analysis of GWAS results from AoU (N = 201,334; African ancestry [AFR]: 50,029; admixed American ancestry (AMR): 36,634; East Asian ancestry [EAS]: 5,083; EUR: 107,415; South Asian ancestry [SAS]: 2,173) and the FinnGen biobank Release 12 (N = 418,856 EUR participants; 56,552 cases and 362,304 controls), totaling 620,190 individuals (84,577 cases and 535,613 controls). The ANX meta-analyses were performed using METAL (Willer et al., 2010) implementing the inverse-variance weighting (IVW) method to optimize precision by weighting effect sizes according to their standard errors while adjusting for ancestry-specific sample sizes. For PAU, we utilized GWAS summary statistics from Zhou et al. (2023) excluding FinnGen data to avoid sample overlap with ANX. The EUR GWAS included 684,355 individuals, while the MA GWAS comprised 1,079,947 individuals of diverse ancestries (AFR: 122,571; AMR: 38,962; EAS: 13,551; EUR: 903,147; SAS: 1,716).

This project was classified by the Uniformed Services University’s Human Research Protections Program Office as research not involving human subjects, following 32 CFR 219.102(e)(1) and applicable DoD policy. The original studies received ethical approval and obtained informed consent from all participants.

### Bidirectional Two-Sample Mendelian Randomization Analysis

We used GWAS summary statistics for PAU and ANX and a bidirectional two-sample MR analysis to assess the relationship between these traits (Hartwig et al., 2016). Independent instrumental variables were selected for PAU and ANX using single nucleotide polymorphisms (SNPs) significantly associated with each trait at a genome-wide significance threshold (*p* < 5×10⁻□). To ensure instrument validity and independence, we applied linkage disequilibrium (LD) clumping (r² < 0.01, window size 10,000 kb). Independent SNP selection, LD clumping, and harmonization were performed using the publicly available EUR reference panel from the 1000 Genomes Project. Palindromic and ambiguous SNPs were excluded to prevent strand mismatches, and harmonization procedures were performed to align effect alleles across exposure and outcome datasets.

Primary causal estimates were derived using Mendelian Randomization Accounting for Pleiotropy and Sample Structure (MR-APSS), which improves causal inference by adaptively selecting valid instruments while accounting for polygenicity and sample structure (Hu et al., 2022). To ensure the robustness of results, we also conducted MR using the IVW method, which provides a weighted regression of SNP-exposure and SNP-outcome associations under the assumption of no directional pleiotropy; weighted median and MR-Egger regression, which detect and correct for pleiotropy; and MR-PRESSO (Mendelian Randomization Pleiotropy RESidual Sum and Outlier) to identify pleiotropic outlier SNPs (Verbanck et al., 2018). All MR analyses were conducted in R using the TwoSampleMR (Hartwig et al., 2016), MRPresso (Verbanck et al., 2018) and MR-APSS packages (Hu et al., 2022).

### Characterizing polygenic overlap

MiXeR analyses were used to determine the shared genetic architecture between ANX and PAU (Frei et al., 2019; Holland et al., 2020). First, univariate MiXeR analyses were conducted to estimate each trait’s polygenicity (the number of genetic variants potentially explaining 90% of SNP heritability) and discoverability (average effect size of causal variants). Prior to bivariate analysis, the adequacy of model power was verified based on finite Akaike information criterion (AIC) scores. Bivariate MiXeR models were then used to estimate the number of unique, overlapping causal variants and the proportion of causal variants whose effect directions were concordant for the two traits. The proportion of polygenic overlap, or Dice coefficient, was also calculated. Conditional Q-Q plots visualized cross-trait enrichment and p-value distributions for the primary phenotype across three significance strata (p ≤ 0.1, 0.01, and 0.001) for association with the secondary phenotype. Analyses examining the shared genetic architecture of ANX and PAU with height were conducted as a control condition. The heritability and genetic correlations were calculated for all traits using LD Score regression (LDSC) v1.0.1.

### Local (Co)Variant Analysis (LAVA)

Local heritability (*h*^2^_SNP_) and genetic correlations (*r_g_*) between PAU and ANX were evaluated using LAVA across 2,495 LD blocks of comparable size (Werme et al., 2022). The false discovery rate (FDR) correction was used to adjust the significance of local *r_g_* values. We did not conduct MA LAVA analyses because validated non-European LD reference panels for LAVA from UKB were unavailable (Bulik-Sullivan et al., 2015).

### ConjFDR Analysis

We used conjFDR analysis to identify loci significantly associated with both phenotypes. CondFDR values were first calculated for the first phenotype (i.e. PAU) conditioned on the second phenotype’s (i.e. ANX) SNP associations, and then repeated using the reverse order, conditioning the second phenotype’s SNP associations on the first (Smeland et al., 2020). The highest of the two condFDR estimates was set as the conjFDR value, ensuring a conservative threshold for identifying joint associations. Associations were deemed significant when the conjFDR value was < 0.05.

### Genomic loci definition and gene expression analysis

SNPs with significant conjFDR values were processed through Functional Mapping and Annotation (FUMA) v1.5.2 to find LD-independent loci (Watanabe et al., 2017). Independent significant SNPs were identified using the EUR 1000 Genomes reference panel and LD block distance criteria (< 250 kb, r2 < 0.6). For each locus, we chose only lead SNPs (r2 < 0.1) with the lowest conjFDR values. ConjFDR-significant lead SNPs that were not genome-wide significant (p < 5x10-8) in the PAU and ANX GWAS were considered novel. Lead SNPs were mapped to genes based on their genomic location, specifically by overlap within gene boundaries or proximity to the nearest gene. BiomaRt (Durinck et al., 2009) and ANNOVAR (Wang et al., 2010) was used to map the nearest gene, when the two disagreed, we verified coordinates in dbSNP (https://www.ncbi.nlm.nih.gov/snp/). Combined Annotation Dependent Depletion (CADD) scores were obtained from OpenTargets (https://genetics.opentargets.org/) (Buniello et al., 2025). Gene expression and tissue specificity analyses were performed in FUMA using Genotype-Tissue Expression (GTEx), with expression levels normalized as transcripts per million to account for differences in library size and sequencing depth. Multiple testing corrections for the enrichment analyses were applied using FDR adjustment.

### FUMA Cell-Type Specific Expression Analysis

To explore cell-type specific associations of PAU and ANX, MAGMA gene-set analyses were conducted using FUMA (Kim et al., 2023). MAGMA gene-level association results from PAU and ANX GWAS summary statistics were uploaded to the FUMA cell-type analysis module. We selected the human brain single-cell RNA sequencing dataset by Siletti et al., (2023), which encompasses transcriptional profiles from various brain regions. MAGMA cell type enrichment analyses assessed whether PAU- and ANX-associated genes were disproportionately expressed in specific cell types. Bonferroni correction was applied across all tested cell types, and conditional analyses were performed within and across datasets to identify cell types most relevant to the shared genetic underpinnings of PAU and ANX.

### Drug repurposing

To identify potential therapeutic targets, druggability, and drug-protein interactions, we queried each locus that was identified from conjFDR through the Target Central Resource Database (TCRD; https://pharos.nih.gov/). TCRD classifies targets based on druggability: Tclin (approved drugs with known mechanisms), Tchem (drug activity meeting specific thresholds), Tbio (biological targets with no known drugs but with Gene Ontology (GO) leaf term annotations or Online Mendelian Inheritance in Man (OMIM) phenotypes, or meeting at least two of the following three conditions: a fractional PubMed count >5, >3 National Center for Biotechnology Gene Reference Intro Function annotations, or >50 commercial antibodies), and Tdark (proteins manually curated in UniProt, but lacking substantial research or druggability criteria). For targets classified as Tclin in TCRD, we queried DrugBank (https://go.drugbank.com) to characterize Click or tap here to enter text.Click or tap here to enter text.the identified drugs and their therapeutic indications.

### Associations of significant shared loci with other phenotypes (PheWAS)

We performed SNP-level phenome-wide association study (PheWAS) using the GWAS ATLAS (Watanabe et al., 2019) to examine loci that were significantly jointly associated with PAU and ANX. For each lead SNP, we reported associations with traits that were significant at p < 5 × 10^-8^.

## Results

### Bidirectional Two-Sample Mendelian Randomization Results

Using the EUR GWAS summary statistics, we observed significant bidirectional causal effects. MR-APSS provided evidence of a significant causal effect of ANX on PAU (OR = 2.33, 95% CI: 1.09–4.98, *p* = 0.029), with pleiotropy (Tau² = 2.73 × 10□¹□) well below typical empirical thresholds (e.g., Tau² < 0.001), indicating minimal violation of instrumental variable assumptions. This was corroborated by the IVW and weighted median methods (Figure 1). The causal effect of ANX on PAU for MR-Egger did not reach statistical significance (*p* > 0.05; Figure 1). However, the MR-Egger intercept indicated no evidence of directional pleiotropy (intercept = 0.0053, *p =* 0.31), and MR-PRESSO did not detect any significant outlying or pleiotropic variant, consistent with results from MR-APSS (Figure 1).

**Figure 1.**
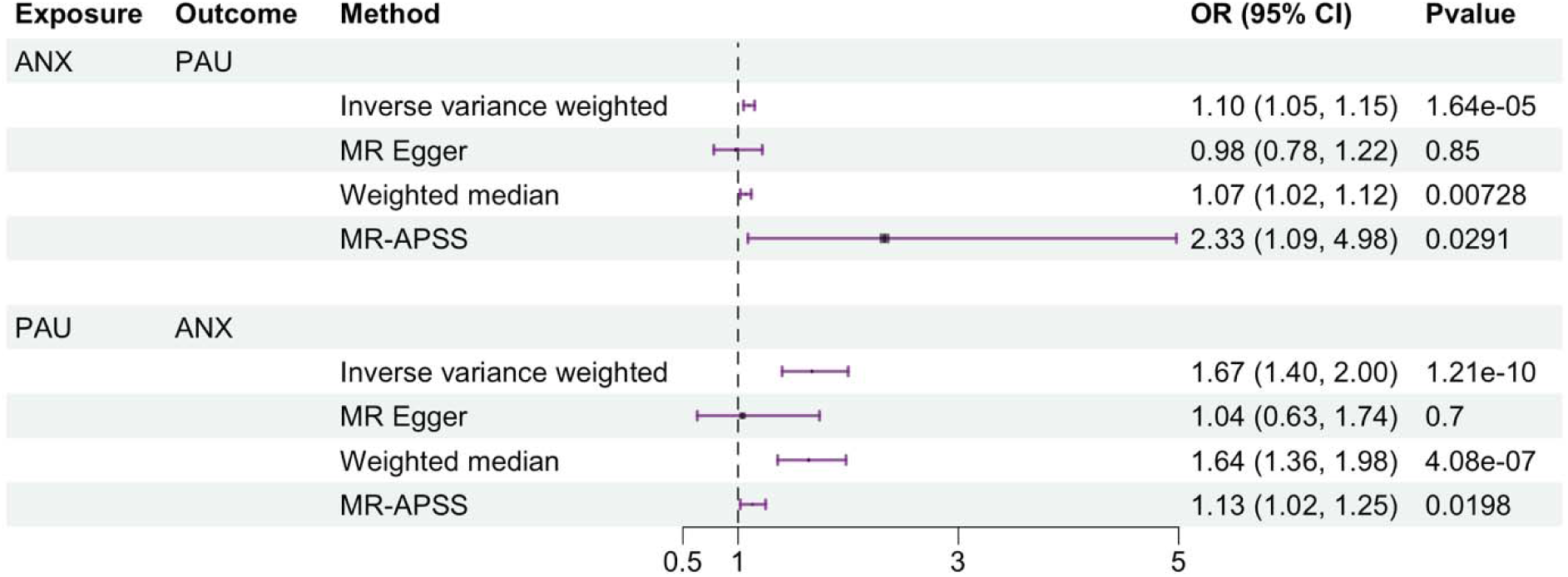
Forest plot summarizing MR analyses of the bidirectional causal relationships between PAU and ANX in EUR individuals. The plot displays odds ratios (OR) with 95% confidence intervals (CI) derived from four MR methods: inverse variance weighted (IVW), MR-Egger, weighted median, and MR-APSS. The top panel illustrates results with ANX as the exposure and PAU as the outcome, while the bottom half represents results with PAU as the exposure and ANX as the outcome.

MR-APSS revealed a significant causal effect of PAU on ANX (OR = 1.13, 95% CI: 1.09–1.18, p = 1.98 × 10□²) with minimal evidence of pleiotropy (Tau² = 1.03 × 10□□; Figure 1). The IVW and weighted median methods also showed a significant causal effect of PAU on ANX (Figure 1). However, the effect was not significant for MR-Egger (OR = 0.99, 95% CI: 0.58–1.69, p = 0.97), and the Egger intercept (0.0096, p = 0.035) suggests potential directional pleiotropy. MR-PRESSO identified one outlier variant (Table S1), and its global test for horizontal pleiotropy was significant (RSS_obs_= 210.61, *p* < 6.67 × 10□□). However, the distortion test showed that removal of the outlier had minimal impact (Distortion *p* = 0.73), and IVW (OR = 1.67, 95% CI: 1.40–2.00, p = 1.99 × 10□□) and weighted median estimates (OR = 1.64, 95% CI: 1.36–1.98, p = 3.25 × 10□□) remained consistent after exclusion, reinforcing the validity of these findings.

Consistent results were obtained using MA GWAS summary statistics. MR-APSS identified a significant causal effect of ANX on PAU (OR = 2.33, 95% CI: 1.08–5.00, p = 0.031; Tau² = 3.61 × 10□¹□). IVW also indicated a significant positive association (Figure S2 and Table S2). However, the weighted median approach and MR-Egger regression and intercept were not significant (p’s > 0.05; Figure S2 and Table S2). MR-PRESSO detected no evidence of heterogeneity or pleiotropy. MR-APSS supported the causal influence of PAU on ANX (OR = 1.20, 95% CI: 1.10–1.30, p = 3.91 × 10□□; Tau² = 4.58 × 10□□; Figure S2). IVW and median weighted analysis also demonstrated a significant causal effect of PAU on ANX (Figure S2 and Table S2). MR-Egger regression was not significant (OR = 1.10, 95% CI: 0.68–1.78, p = 0.699), however, the MR-Egger intercept (0.0073, p = 0.0477) indicated marginal evidence of directional pleiotropy. MR-PRESSO detected no outliers, suggesting no substantial heterogeneity or horizontal pleiotropy.

### Shared genomic architecture (MiXeR)

Univariate MiXeR models demonstrated sufficient power for bivariate analyses, as all models yielded finite AIC values. MiXeR analysis identified significant polygenic overlap between PAU and ANX. In the EUR cohort, the traits were moderately genetically correlated (*r_g_* = 0.44), with moderate polygenic overlap (Dice coefficient = 52.5%). Specifically, of the 8,395 causal variants estimated to be linked to PAU and the 9,453 linked to ANX, an estimated 4,698 were shared, reflecting substantial genetic commonality between these two phenotypes. Additionally, 86.4% of the shared estimated causal variants were concordant for effect direction (Table S3).

MiXeR analysis using MA GWAS summary statistics revealed a moderate genetic correlation between PAU and ANX (*r_g_* = 0.44) and moderate polygenic overlap (Dice coefficient = 56.3%). Of the estimated 8,378 causal variants for PAU and 8,622 for ANX, 4,790 variants were shared. Moreover, 81.9% of the shared variants exhibited concordant effect directions, further supporting their genetic interrelationship (Table S3).

Conditional Q-Q plots illustrated enrichment patterns, with PAU-associated SNPs increasingly enriched within the top significance strata for ANX, and similarly, ANX-associated SNPs were enriched among the top PAU-associated strata (Figure 2).

**Figure 2.**
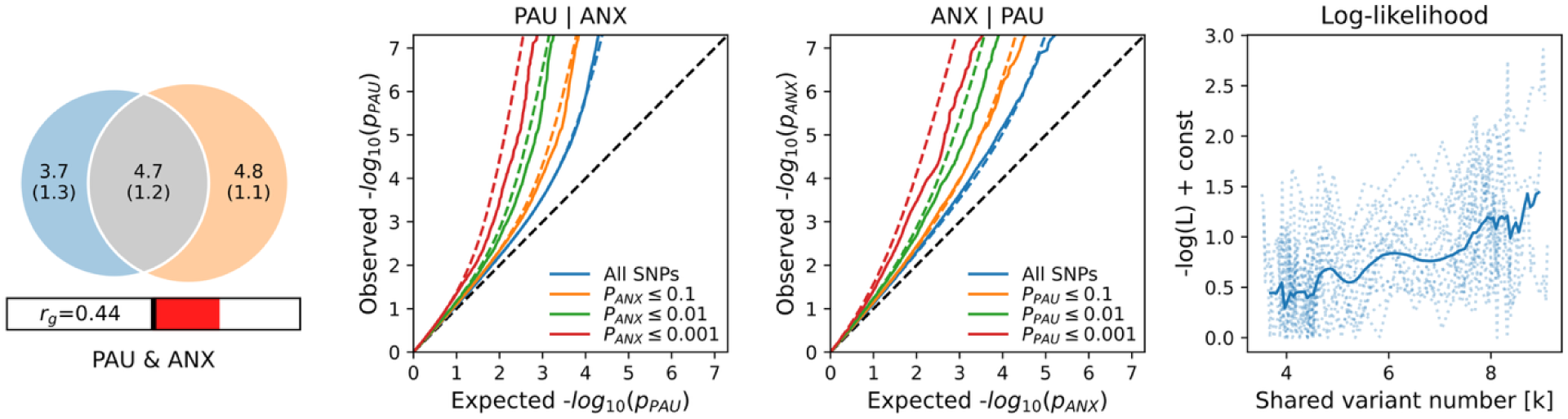
PAU-ANX bivariate MiXeR results in EUR individuals. The Venn diagram (left) illustrates the estimated numbers of causal genetic variants (in thousands) unique to PAU (blue), unique to ANX (orange), and variants shared between both traits (gray). Genetic correlation (r_g_) is indicated beneath. The conditional Q–Q plots (middle) demonstrate cross-trait enrichment, showing increased polygenic signals for PAU conditional on ANX (PAU|ANX) and for ANX conditional on PAU (ANX|PAU), across varying thresholds of SNP significance (p ≤ 0.1, 0.01, 0.001). The right panel shows the log-likelihood curve used to estimate the number of shared causal variants, illustrating model fit across variant number estimations.

To validate our findings, we conducted a control analysis examining genetic overlap between PAU and height, identifying minimal genetic overlap with PAU (11.2%). Specifically, of the approximately 8,390 causal variants linked to PAU and the 3,997 linked to height, only 670 were shared. The genetic correlation was near-zero (*r_g_* = –0.044), with low concordance between the shared causal variants (33.0%; Table S3).

### Local (Co)Variant Analysis

LAVA examined 2,495 independent genomic regions, of which 1,578 showed significant local heritability (p < 0.05) for both PAU and ANX. Local genetic correlations between PAU and ANX were significant in 20 regions after FDR correction (q < 0.05), with 70% of these (14/20) exhibiting positive correlations (Table S4).

### Shared genetic loci (cond/conjFDR)

At conjFDR < 0.05, we identified 97 loci associated with both PAU and ANX in the EUR summary statistics. Among these, 90 were not genome-wide significant in the PAU summary statistics, and 93 were not significant in the ANX summary statistics. Eighty-nine lead SNPs (91.8%) had concordant effect directions for PAU and ANX, and 8 had discordant effect directions (Table S5). The 97 loci mapped to 97 genes, including *DRD2* and *PDE4B*, which have known roles in dopaminergic and cyclic AMP (cAMP) signaling pathways, respectively.

In the MA cohort, we identified 92 loci jointly associated with PAU and ANX (Figure S3), of which 85 and 89 variants were not genome-wide significant in the PAU and ANX summary statistics, respectively. Among the lead SNPs, 84 (91.3%) showed concordant effect directions between PAU and ANX, while 8 (8.7%) were discordant. Comparing the MA to the EA results (Table S6), 28 SNPs (highlighted in red, column “rsID”) and 41 mapped genes (highlighted in green, column “Gene”) overlapped. Thus, three quarters (69/92) of genes were overlapping across EUR and MA analyses.

### FUMA genotype-tissue expression analysis

Analysis of GTEx data revealed that genes linked to the 89 concordant lead SNPs in EUR individuals were significantly differentially expressed across multiple tissues, including brain, nerve, stomach, colon, and adrenal gland (Figure 3A). In the 54-tissue analysis, significant enrichment was identified in key brain regions implicated in psychiatric disorders, particularly the basal ganglia, frontal cortex, hypothlamaus, anterior cingulate cortex, hippocampus, amygdala, and hypothalamus (Figure 3B). In contrast, genes linked to the 8 non-concordant SNPs showed no significant differential expression across tissues.

**Figure 3.**
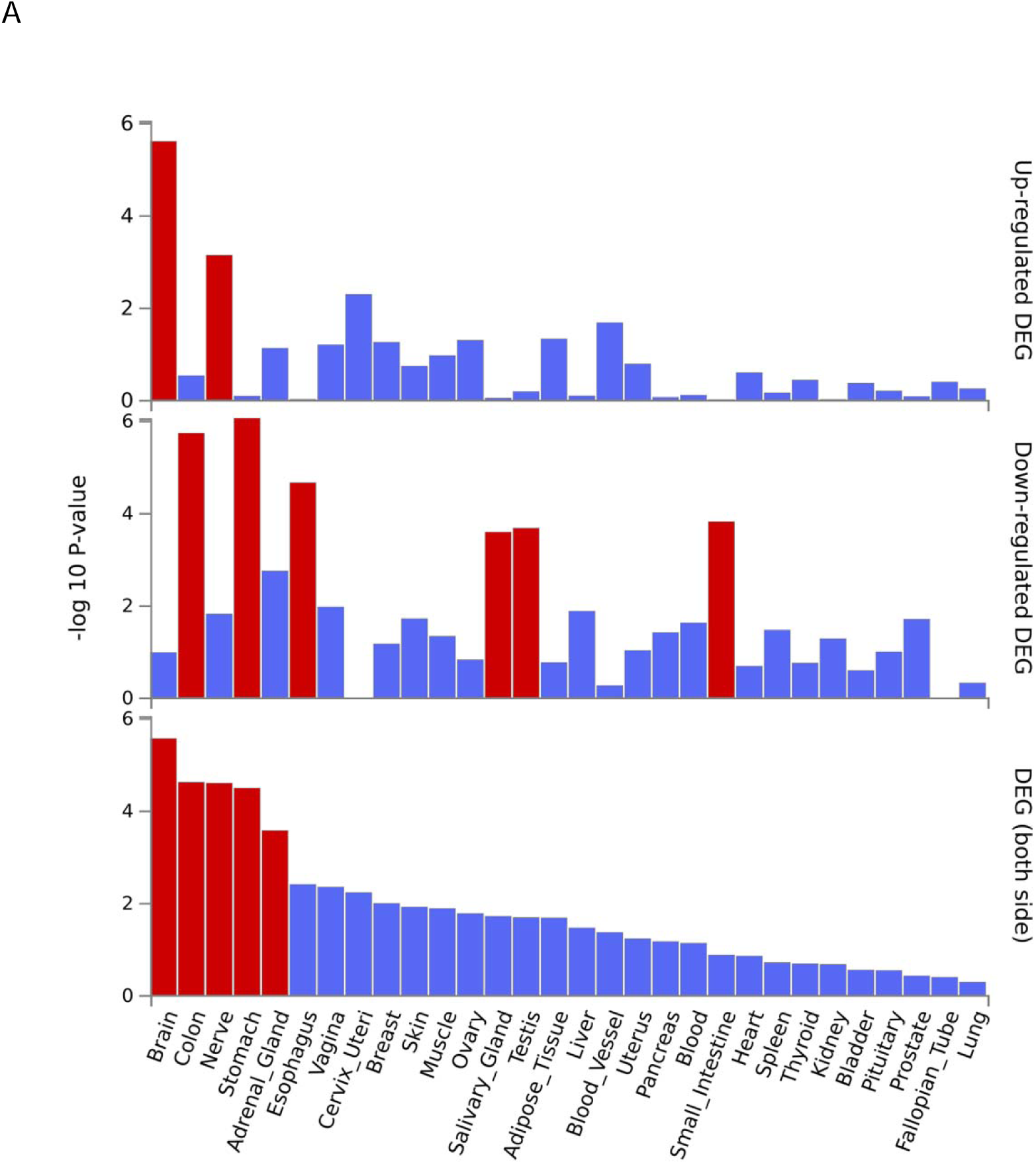
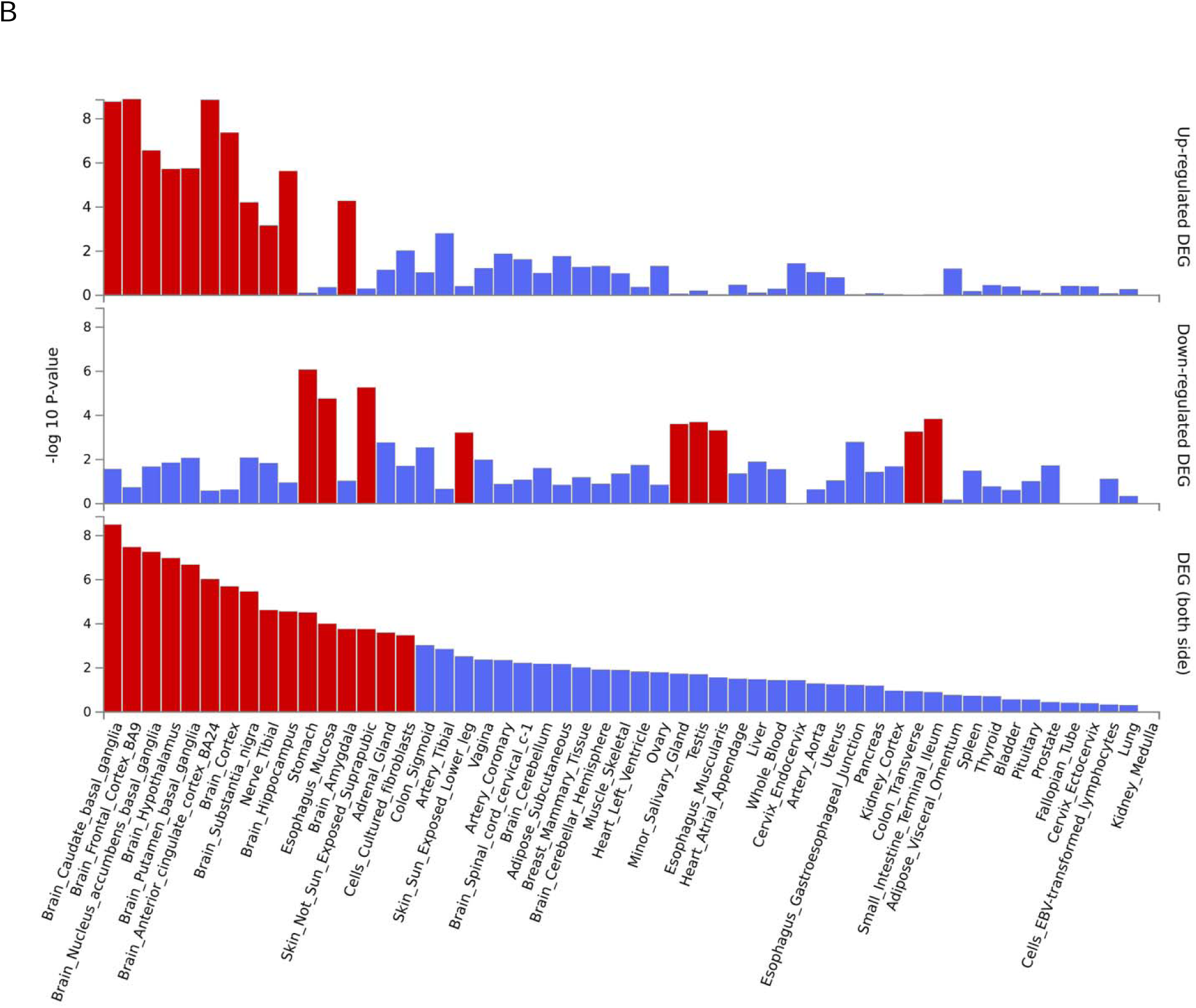
GTEx v8 tissue-specific differential gene expression for genes linked to concordant SNPs in EUR individuals. The analysis shows significant differential expression (DEG) across 30 general tissue types (A) and 54 tissue types (B). The top panel illustrates tissues with genes significantly up-regulated (red bars); the middle panel highlights tissues with significantly down-regulated genes (blue bars). The bottom panel integrates both up- and down-regulated DEGs across all tissues, with brain tissues prominently enriched (highlighted in red), emphasizing their potential role in psychiatric disorders related to PAU and ANX.

Genes linked to the 84 concordant lead SNPs in MA individuals were significantly differentially expressed across the brain, stomach, colon, esophagus, and adrenal gland (Figure S4A). In the 54-tissue analysis, significant enrichment was identified in a similar set of key brain regions as the EUR analysis (Figure S4B). Genes linked to the 8 non-concordant SNPs showed no significant differential expression across tissues.

### FUMA cell type enrichment analysis

Using MAGMA, we identified 207 significant cell-type associations across brain regions that were implicated in the shared genetic architecture of PAU and ANX. These associations spanned 10 brain regions, 118 subregions or clusters, and 13 neuronal cell types, based on single-cell RNA sequencing data from Siletti et al. (2023). The most significant cell-type enrichment was in neurons and oligodendrocytes in the thalamus (PoN.MG). Neuron and oligodendrocyte enrichment was identified across several brain subregions, namely subregions of the myelencephalon, cerebral cortex, pons, midbrain, hippocampus. suggesting that these areas are potentially critical neural substrates for the pathophysiology of PAU and ANX (Figure 4, Figure S5, and Table S7).

**Figure 4.**
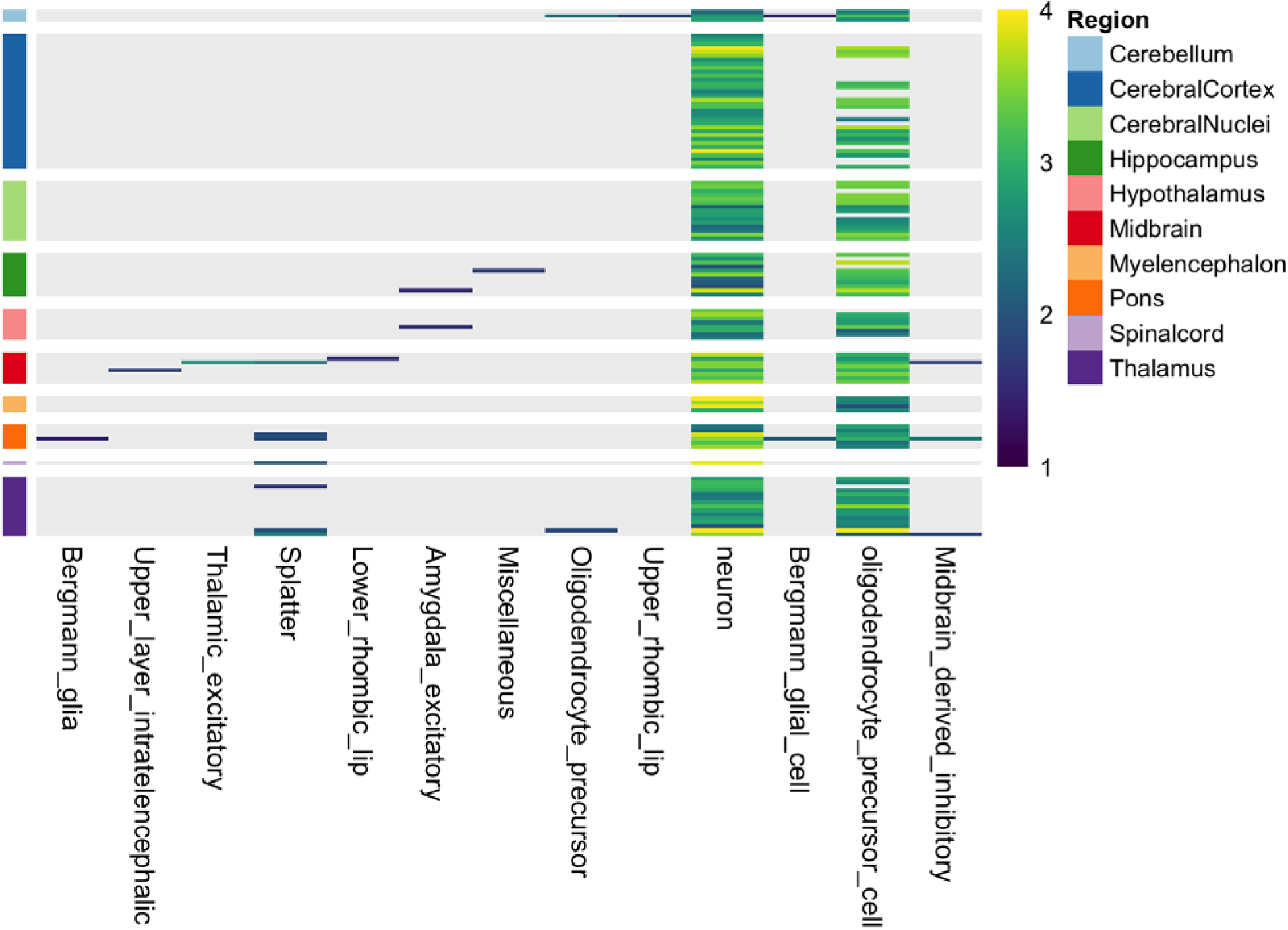
Heatmap of cell-type-specific enrichment across brain subregions for PAU and ANX in EUR individuals. This heatmap depicts row-scaled average enrichment signals across brain subregions. and various cell types (columns, hierarchically clustered with a top dendrogram). White lines separate regional blocks. Enrichment scores represent the mean of – log□□(P) values for significant cell-type associations identified using MAGMA gene-set analyses of PAU- and ANX-associated genes. Rows are grouped into 10 major anatomical brain regions, indicated by a colored annotation bar and corresponding legend (see Table S7 and Figure S5 for additional detail). Region classifications follow Siletti et al. (2023), mapping subregions to higher-order brain structures. Yellow indicates stronger enrichment (i.e., greater association of disease-related gene expression with that cell type in the subregion), and purple indicates weaker enrichment.

### Drug repurposing

Among the genes identified in EUR and MA individuals, four—*PDE4B*, *DRD2*, *TYR*, and *KCNA4—*were classified as Tclin targets, indicating that they are involved in the mechanisms of action for FDA-approved drugs (Tables S5, S6, and S8). *PDE4B*, a key regulator of cAMP signaling involved in neuroinflammation and psychiatric disorders, is associated with several pharmacological agents commonly used in respiratory conditions like asthma and chronic obstructive pulmonary disease. It is important to note that identification of gene–drug interactions does not imply that all linked compounds would be therapeutic. For example, *PDE4B* is a target of both anti-inflammatory agents (e.g., roflumilast) and stimulants (e.g., theophylline), the latter of which are known to exacerbate anxiety in some individuals. *DRD2*, encoding the dopamine D2 receptor, central to reward processing and psychopathology, was linked to over 80 approved drugs, including antipsychotics, antidepressants, dopamine agonists, and adrenergic modulators. Given the D2 receptor*’*s established role in anxiety and addictive behaviors, dopaminergic agents such as risperidone (used off-label for treating anxiety), trifluoperazine (approved for treating anxiety), and maprotiline (indicated for treating depression with anxiety) may present repurposing opportunities worthy of further study for treating PAU-ANX comorbidity. *TYR*, targeted mainly by dermatologic and metabolic agents, and *KCNA4*, a voltage-gated potassium channel linked to modulators used in neurological, neuromuscular, and cardiovascular conditions (e.g., multiple sclerosis, Lambert–Eaton myasthenic syndrome, arrhythmias, hypertension), represent additional Tclin targets, though their direct relevance to PAU and ANX biology is less clear.

### Associations of significant shared loci with other phenotypes (PheWAS)

We examined whether the 97 shared genetic loci of PAU and ANX identified in the EUR cohort were associated with other phenotypes in the GWAS ATLAS database. Approximately half of the SNPs (49 loci; 50.5%) were previously associated with other phenotypes (Table S9). Of these, 26 SNPs were associated with psychiatric traits, including schizophrenia (9 loci), depression (11 loci), and neuroticism (6 loci). Twenty-one SNPs were associated with metabolic traits, such as body fat or BMI (16 loci).

Similar results were found for the 92 shared genetic loci from the MA cohort. Forty-seven loci (51.1%) were previously associated with other phenotypes. Of these, the most common phenotypes were psychiatric (26 loci), such as schizophrenia (7 loci), depression (10 loci), and neuroticism (7 loci). Seventeen SNPs were also associated with metabolic traits, 14 of which were associated with body fat or BMI (Table S10).

## Discussion

This study elucidated the shared genetic architecture and underlying neurobiological mechanisms of PAU and ANX, two conditions that frequently co-occur. Using complementary methods (MR, MiXeR, LAVA, and conjFDR), we identified reciprocal causal relationships, substantial polygenic overlap, and specific shared genetic loci. These findings extend beyond genetic correlation by pinpointing the specific variants, pathways, and tissues underlying comorbidity.

We identified bidirectional causal effects between PAU and ANX across most MR methods in both the EUR and MA cohorts. MR-Egger, which did not yield any significant results, is characterized by lower statistical power due to its accounting for pleiotropy (Hu et al., 2022). The wide CI observed in the MR-APSS analysis of ANX on PAU likely reflects weaker instrument strength, due to the lower heritability of the ANX GWAS and fewer robust genetic associations (Hu et al., 2022). While MR cannot fully exclude pleiotropy, the convergence of results across methods supports a reciprocal relationship. MiXeR demonstrated moderate polygenic overlap, with the majority of shared variants showing concordant effects, while LAVA pinpointed genomic regions with significant local correlations. ConjFDR further identified nearly 100 loci jointly associated with both phenotypes. Taken together, these findings extend prior genetic studies (Ahn et al., 2025; Colbert et al., 2021) by clarifying the presence and directionality of shared variants in the largest combined analysis of PAU and ANX to date.

The neurobiological interpretation of these loci converges on brain regions and signaling pathways implicated in addiction and anxiety. Genes such as *DRD2* and *PDE4B* highlight dopaminergic and cAMP signaling, respectively, while enrichment analyses implicated the prefrontal cortex, anterior cingulate cortex, hippocampus, hypothalamus, amygdala. Cell-type analyses further revealed strong enrichment in cortical, hippocampal, midbrain, and thalamic neurons—regions and circuits central to cognitive control, stress regulation, and reward processing. Notably, our *DRD2* finding aligns with animal models showing that anxiety-prone mice exhibit striatal dopamine receptor imbalance that enhances the anxiolytic and reinforcing effects of alcohol (Bocarsly et al., 2024), underscoring dopaminergic dysfunction as a shared mechanism underlying ANX–PAU comorbidity.

Our analyses provide important insights into traits that share genetic risk with PAU and ANX. Many loci that confer risk for PAU and ANX are associated with other psychiatric traits, including depression, neuroticism, and schizophrenia, consistent with prior studies (Polimanti et al., 2019; Ward et al., 2017; Wiström et al., 2022). Thus, genetic pleiotropy representing general psychopathological risk may play a role in the shared etiological processes underlying these disorders (Chen et al., 2025; Garey et al., 2020). Additionally, shared genetic risk was also associated with metabolic phenotypes such as body fat and BMI, consistent with our prior work linking AUD and BMI (Malone et al., 2025). These overlaps suggest that medications targeting neuroendocrine pathways involved in appetite regulation, such as ghrelin and glucagon-like peptide-1 receptor agonists, may also hold promise for individuals with PAU and ANX (De Giorgi et al., 2025; Farokhnia et al., 2025).

A key limitation of the present work is that the MA analyses are dominated by individuals of EUR ancestry. Thus, the observed consistency in our results between the two cohorts may reflect the strong influence of the EUR sample. Additionally, the bidirectional causal associations between PAU and ANX, revealed through MR analyses, require careful consideration of the underlying assumptions, particularly the potential for residual pleiotropy. Finally, our findings should be interpreted in light of heterogeneous case definitions and variable ascertainment across contributing cohorts, which can dilute phenotypes and inflate broad cross-trait sharing.

Future research should prioritize functional genomic investigations to elucidate the precise biological mechanisms underlying the identified concordant loci. Animal models can offer critical insights into gene-environment interactions and neurodevelopmental processes that drive the bidirectional effects between these traits (Anker & Kushner, 2019). Longitudinal studies investigating the temporal dynamics of the PAU-ANX relationship across the lifespan are also warranted, as the bidirectional effects between PAU and ANX may differ across developmental stages (D’Aquino et al., 2024; Ummels et al., 2022).

In conclusion, we found consistent evidence of substantial genetic overlap and bidirectional causal effects between PAU and ANX, underscored by shared genetic loci enriched in neurobiologically relevant brain regions. The shared loci and pathways converge on neurobiological systems that regulate stress, reward, and emotion, offering new avenues for mechanistic insight and therapeutic development.

## Data Availability

All data produced in the present work are contained in the manuscript

## Notes

Conflict of Interest Disclosures: Dr. Kranzler is a member of advisory boards for Altimmune and Clearmind Medicine; a consultant to Sobrera Pharmaceuticals, Altimmune, Lilly, and Ribocure; the recipient of research funding and medication supplies for an investigator-initiated study from Alkermes and a company-initiated study from Altimmune; and an inventor on U.S. provisional patent “Multi-ancestry Genome-wide Association Meta-analysis of Buprenorphine Treatment Response.” Dr. Polimanti received a research grant from Alkermes outside the scope of the present study and received an honorarium for his editorial work in the journal Complex Psychiatry.

Funding: Dr. Kranzler is supported by the Veterans Integrated Service Network’s Mental Illness Research, Education and Clinical Center; U.S. Department of Veterans Affairs grant I01 BX004820 and NIAAA grant R01 AA030056. Dr. Gray is supported by NIAAA grant R01 AA030041 and Department of Defense grant HU0001-22-2-0066. Dr. Polimanti is supported by NIMH grant RF1 MH132337. Dr. Davis is supported by the Office of Academic Affiliations, Advanced Fellowship Program in Mental Illness Research and Treatment, Department of Veterans Affairs.

### Competing Interest Statement

Dr. Kranzler is a member of advisory boards for Altimmune and Clearmind Medicine; a consultant to Sobrera Pharmaceuticals, Altimmune, Lilly, and Ribocure; the recipient of research funding and medication supplies for an investigator-initiated study from Alkermes and a company-initiated study from Altimmune; and an inventor on U.S. provisional patent Multi-ancestry Genome-wide Association Meta-analysis of Buprenorphine Treatment Response.
Dr. Polimanti received a research grant from Alkermes outside the scope of the present study and received an honorarium for his editorial work in the journal Complex Psychiatry.

### Funding Statement

This study was funded by:
Dr. Kranzler is supported by the Veterans Integrated Service Network Mental Illness Research Education and Clinical Center; U.S. Department of Veterans Affairs grant I01 BX004820 and NIAAA grant R01 AA030056. Dr. Gray is supported by NIAAA grant R01 AA030041 and Department of Defense grant HU0001-22-2-0066. Dr. Polimanti is supported by NIMH grant RF1 MH132337. Dr. Davis is supported by the Office of Academic Affiliations Advanced Fellowship Program in Mental Illness Research and Treatment Department of Veterans Affairs.

### Author Declarations

The study used ONLY openly available human data that were originally located at:https://www.ebi.ac.uk/gwas/

