## Supplemental Figure for "Shared Genetic Architecture and Neurobiological Pathways of Problematic Alcohol Use and Anxiety Disorders"

**Figure S1. Meta-Analysis Results of ANX GWAS combining AoU and FinnGen cohorts in EUR individuals.** (A) Manhattan plot depicting the genome-wide association results for ANX meta-analysis. The x-axis represents genomic coordinates across chromosomes, and the y-axis shows the −log10 p-values for each SNP. The red horizontal line indicates the genome-wide significance threshold (p < 5 × 10⁻⁸). (B) Quantile-Quantile (QQ) plot illustrating the observed versus expected −log10 p-values under the null hypothesis. The deviation from the diagonal line indicates the presence of true associations beyond what is expected by chance.

A

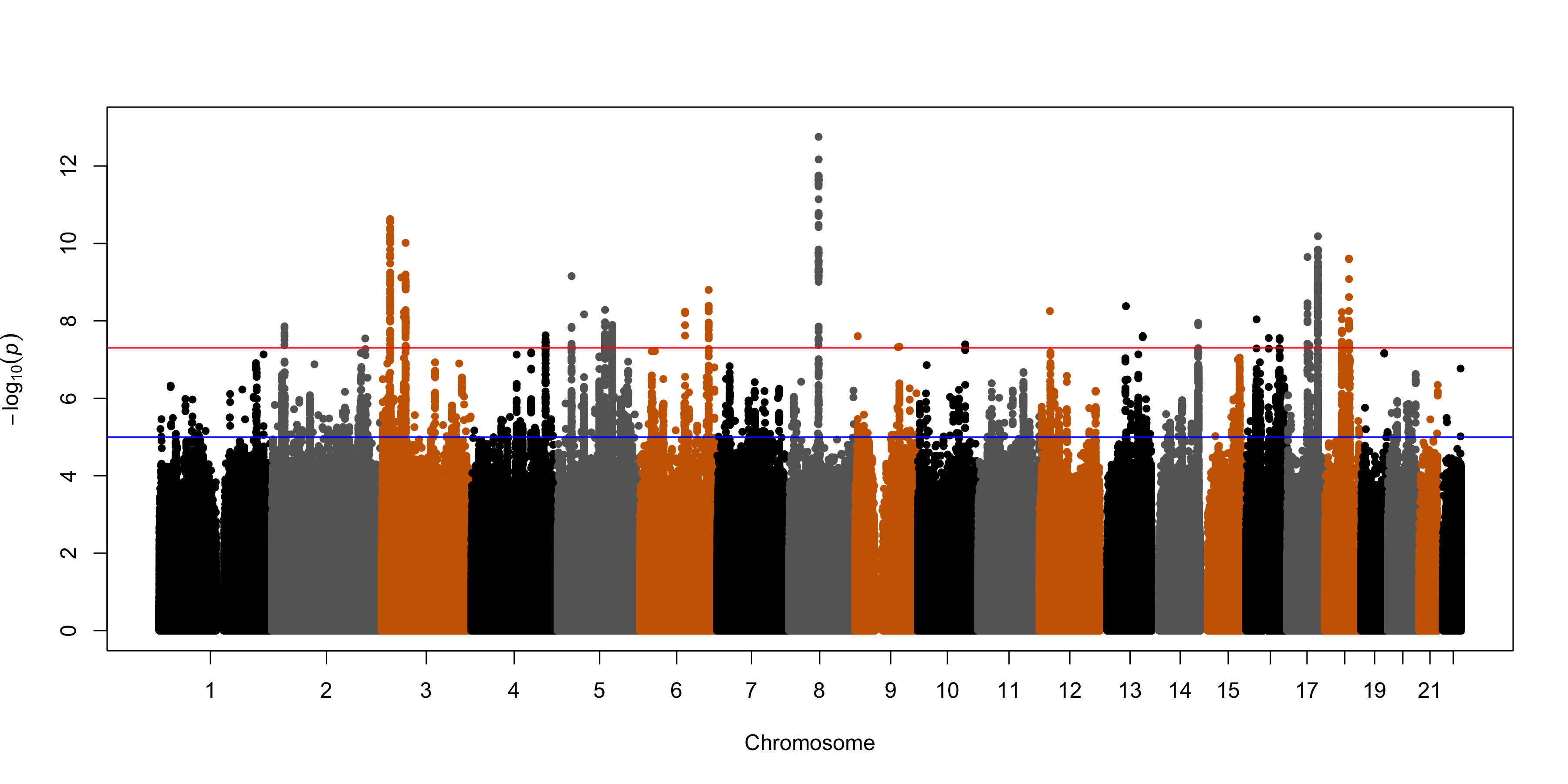

**B**

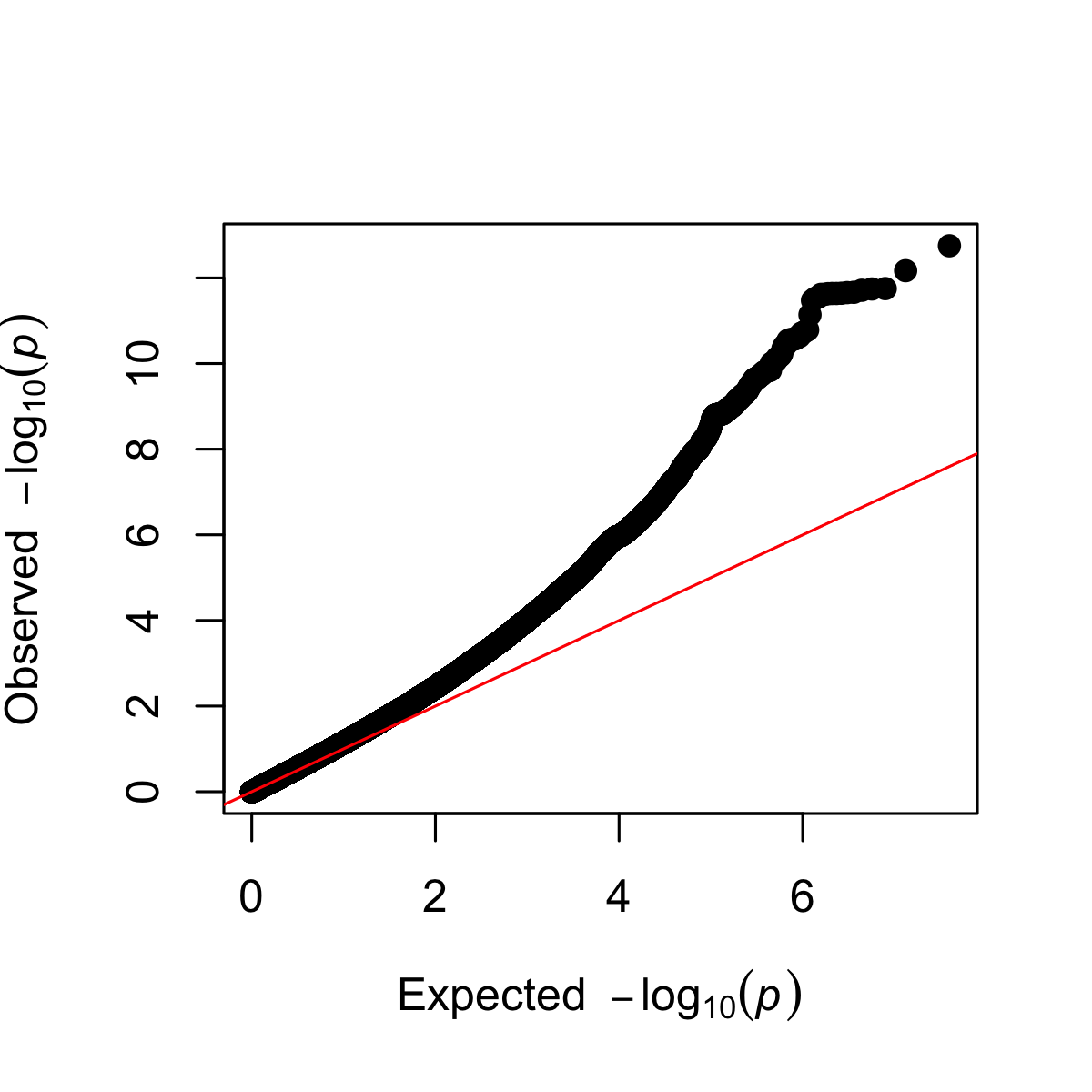

**Figure S2. Forest Plot of MR Results in MA individuals.** Forest plot summarizing MR analyses examining the bidirectional causal relationships between PAU and ANX in individuals of MA. The plot displays odds ratios (OR) with 95% confidence intervals (CI) derived from four MR methods: inverse variance weighted (IVW), MR-Egger, weighted median, and MR-APSS. The top panel illustrates results with ANX as the exposure and PAU as the outcome, while the bottom half represents results with PAU as the exposure and ANX as the outcome.

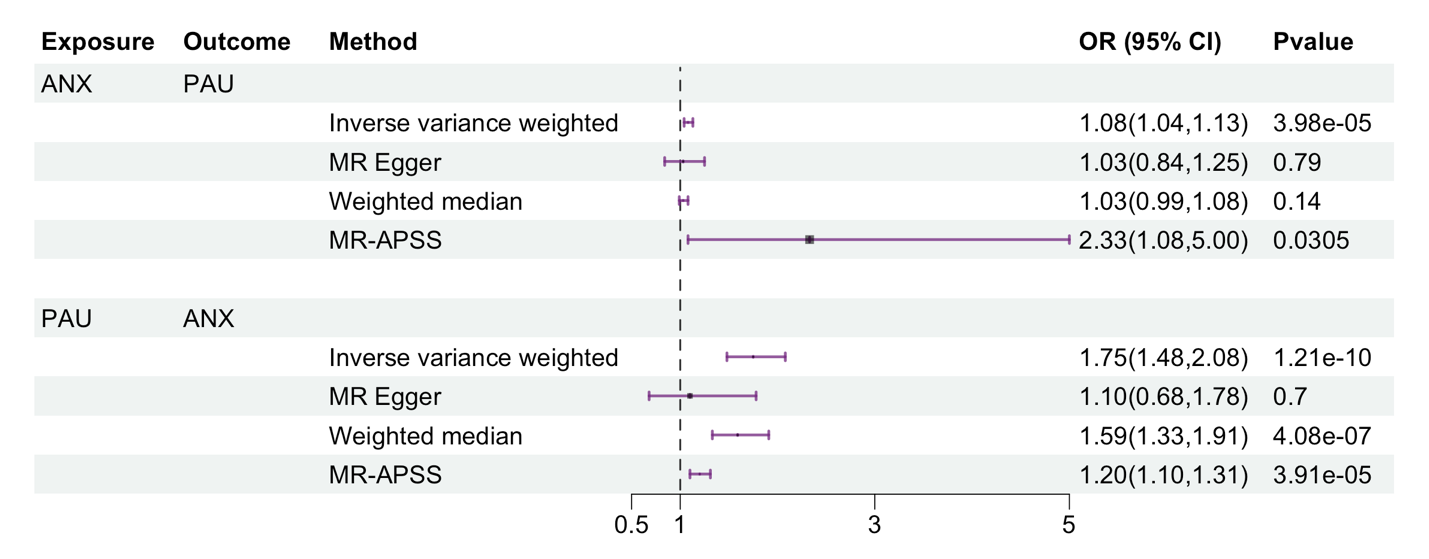

**Figure S3. conjFDR manhattan plot for PAU and ANX in MA individuals.** The plot displays the genomic loci jointly associated with PAU and ANX using the conjFDR approach. Each dot represents a SNP plotted by chromosomal position (x-axis) and the –log10 of the conjFDR-adjusted p-value (y-axis). The blue horizontal line denotes the significance threshold (conjFDR < 0.05). Green-highlighted SNPs indicate lead variants surpassing the significance threshold.

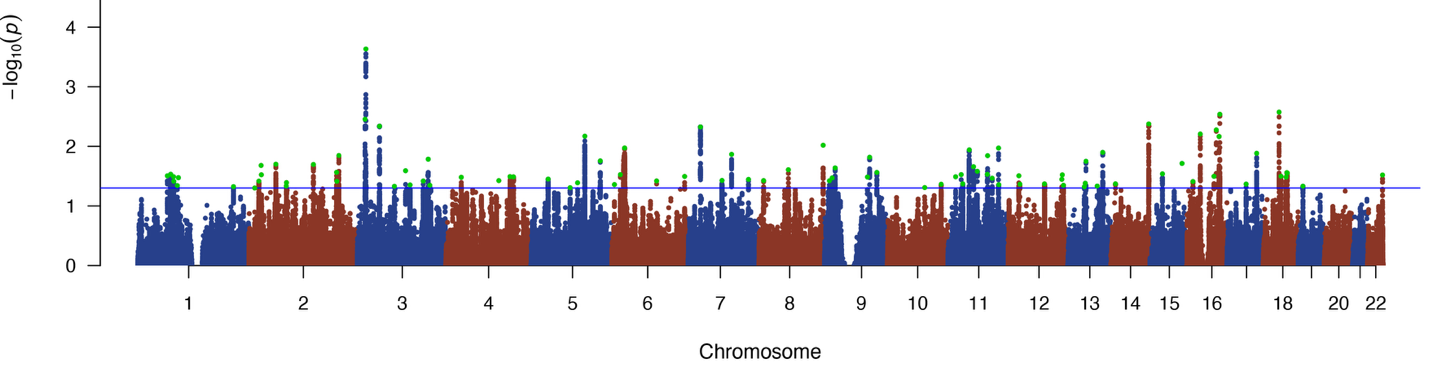

**Figure S4. GTEx v8 tissue-specific differential gene expression for genes linked to concordant SNPs in MA individuals.** The analysis shows significant differential expression (DEG) across 30 general tissue types (A) and 54 tissue types (B). The top panel illustrates tissues with genes significantly up-regulated (red bars); the middle panel highlights tissues with significantly down-regulated genes (blue bars). The bottom panel integrates both up- and down-regulated DEGs across all tissues, with brain tissues prominently enriched (highlighted in red), emphasizing their potential role in psychiatric disorders related to PAU and ANX.

A

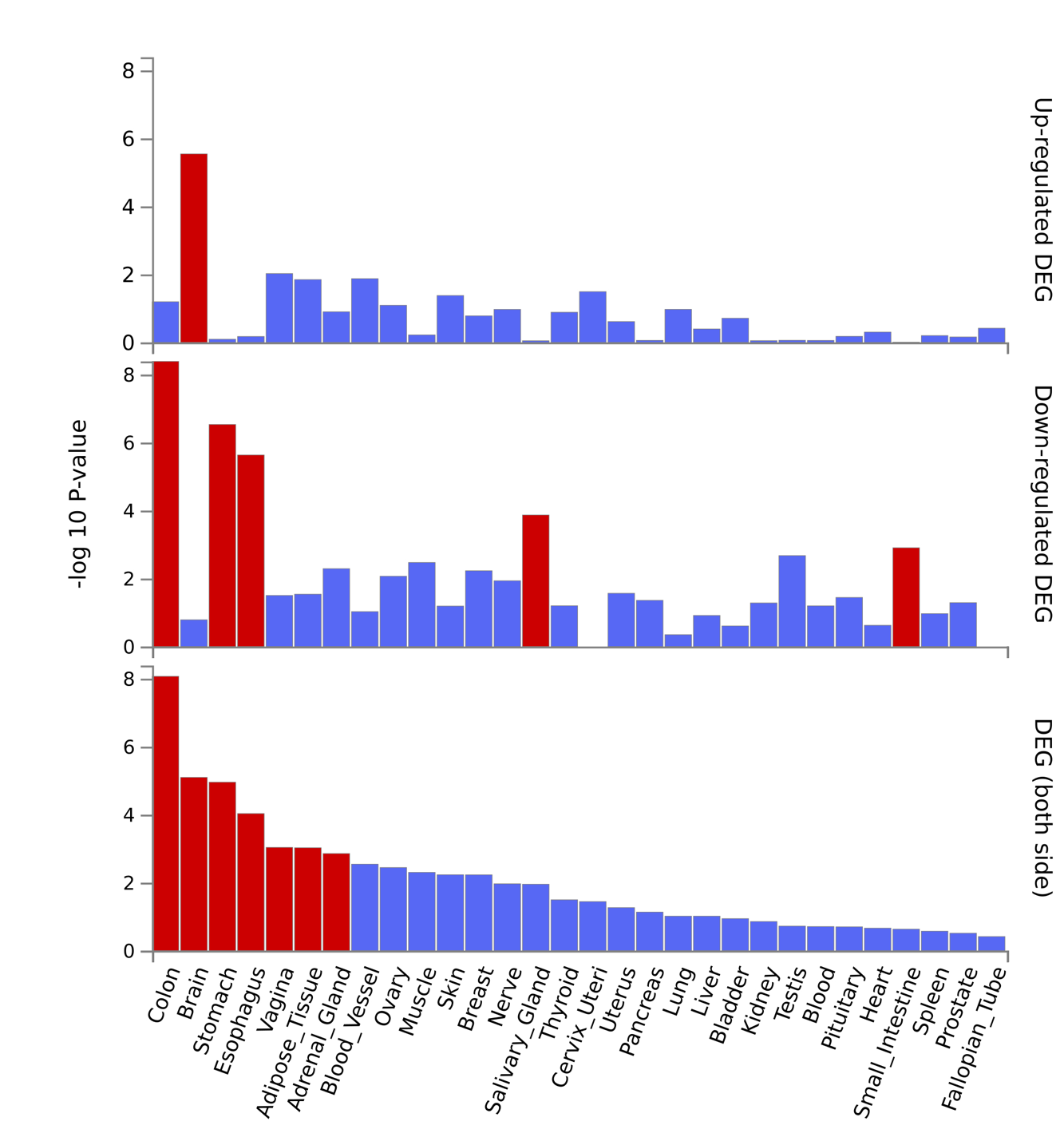

B

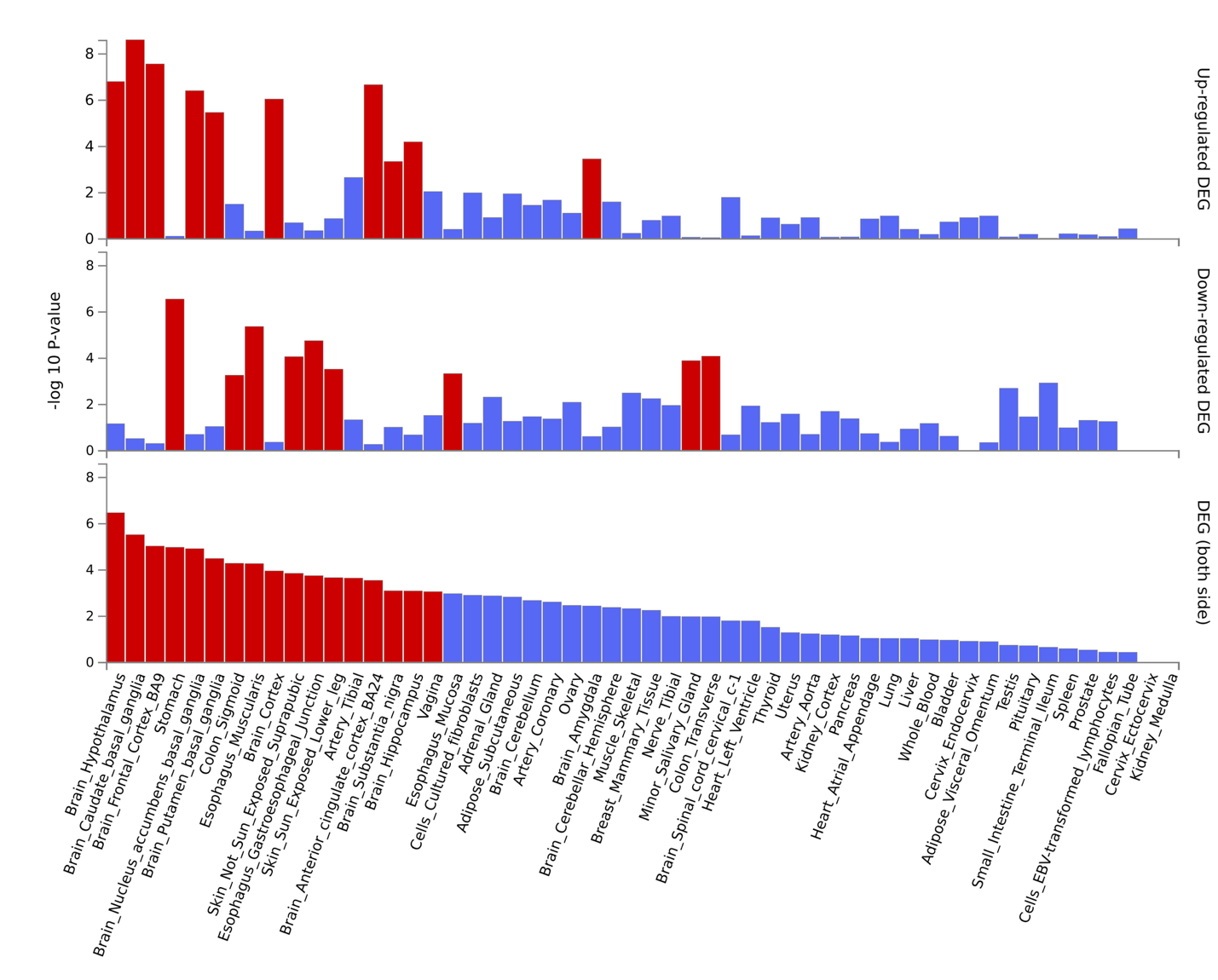

**Figure S5. Cell-Type-Specific Enrichment Across Brain Subregions for PAU and ANX in EUR individuals.** This heatmap depicts row-scaled average enrichment signals (mean –log₁₀(P)) for significant cell-type associations identified from MAGMA gene-set analyses of genes linked to PAU and ANX. Rows represent significant subregions across the human brain. Columns represent cell types hierarchically clustered based on similarity in enrichment profiles. Yellow indicates stronger enrichment (i.e., higher association of disease-related gene expression with a given cell type in a specific subregion), while purple indicates weaker relative enrichment.

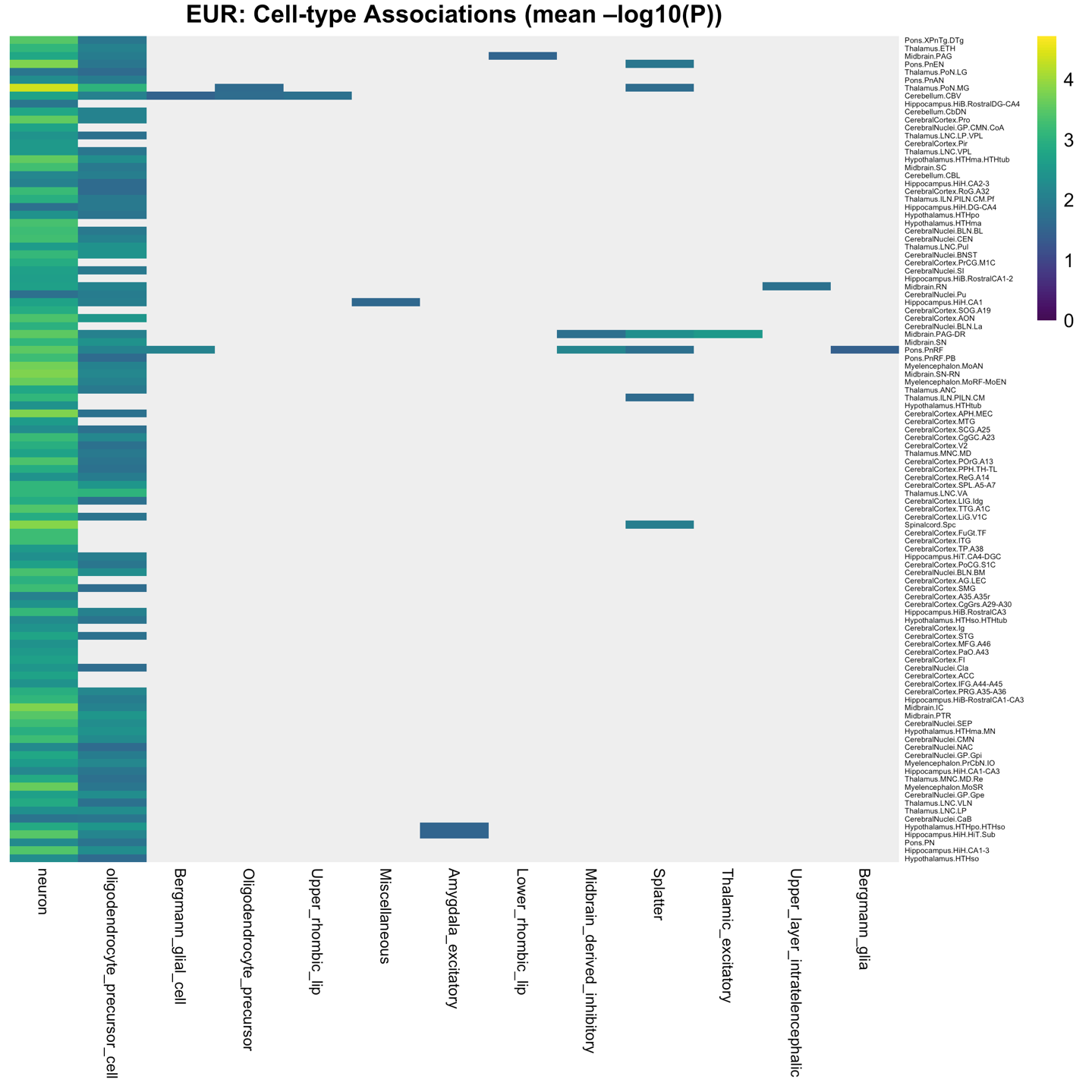
